# Longitudinal trajectories of psychological resilience and cognitive impairment among older adults: evidence from a national cohort study

**DOI:** 10.1101/2024.09.02.24312919

**Authors:** Peicheng Wang, Ruihua Li, Yanhua Chen

## Abstract

**Background:** The relationship between resilience trajectories and cognitive health is not well understood. This study aimed to identify subgroups of psychological resilience trajectories in a national sample of older adults and to examine the association with cognitive impairment over time.

**Methods:** This study used data from the Chinese Longitudinal Healthy Longevity Survey (CLHLS) from 2008 to 2018, and 2,788 respondents were included in this prospective analysis. Using a group□based trajectory modeling approach, we identified resilience trajectory groups within a 6-year period. Cox proportional hazards models were used to assess the relationship between the resilience trajectory groups and cognitive impairment.

**Results:** Three distinct trajectories of psychological resilience, including decreasing resilience group (n=131,7.1%), persistent middle resilience group (n=1,808, 58.1%), and persistent high resilience group (n=849, 34.8%). During the 6-year follow-up, compared to those with persistent high resilience, participants with persistent middle resilience (HR=1.43, 95% CI=1.14-1.79) and decreasing resilience (HR=2.46, 95% CI=1.76-3.43) remained consistent associated with a higher risk of cognitive impairment. The associations between resilience trajectories and cognitive impairment varied by lifestyle and health conditions.

**Conclusions:** Psychological resilience is a relatively stable trait among older adults in China, with most individuals maintaining a persistently high or middle level of resilience throughout the follow-up period; however, declining psychological resilience was significantly associated with the risk of cognitive impairment. Therefore, developing targeted interventions to strengthen psychological resilience in older adults is crucial for promoting cognitive health and successful aging, especially for those who have unhealthy lifestyles and with poorer health conditions.

## Introduction

Cognitive impairment is common among older adults. With the population aging and growing, dementia, the most severe expression of cognitive impairment, has become a severe public health concern and the leading cause of disability in older adults worldwide (Feigin et al., 2019; Livingston et al., 2020). Reducing cognitive impairment is an important way to prevent dementia from developing. In the daily lives of older adults, cultivating healthy lifestyle habits (Jia et al., 2023; Kivipelto et al., 2018) like regular physical activity (Gallardo-Gómez et al., 2022), abstaining from smoking (Anstey et al., 2007) and alcohol consumption (Topiwala et al., 2017), and effectively managing chronic diseases (Vassilaki et al., 2015) and stress reduction (Franks, Rowsthorn, et al., 2023) are essential for preserving cognitive function and lowering the risk of cognitive decline.

Psychological resilience is a personal psychological resource or internalized characteristic, indicating an individual’s ability to adapt in the face of challenges (e.g., adversity, trauma, or stress), which is a critical factor shaping health and well-being in later life (Taylor & Carr, 2021; Ungar & Theron, 2020). Resilient individuals exhibit effective coping strategies, emotional regulation, and problem-solving skills, which can buffer against the negative impact of stress on cognitive function (Harvanek et al., 2021; Machado et al., 2014). While there is growing interest in the association between resilience and cognitive function (Franks, Bransby, et al., 2023; Jiang et al., 2024; McDaniel et al., 2022), few of them have considered the long-term trajectory of resilience, overlooking the fact that resilience is a relatively stable psychological trait as over time (Linnemann et al., 2020; Taylor & Carr, 2021).

To fill this knowledge gap, we captured psychological resilience trajectories and examined the association between the resilience trajectories and cognitive impairment. Additionally, we explore the role of lifestyle and health conditions on these associations, with the aim of contextualizing them within daily life dementia prevention. The findings from this study will contribute to a deeper understanding of the relationship between psychological resilience and cognitive function, informing targeted interventions to promote cognitive health across diverse populations.

## Methods

### Data Source and Sample

Data were drawn from the Chinese Longitudinal Healthy Longevity Survey (CLHLS), a nationally representative prospective cohort study of older participants (Yi, 2008; Zeng, 2012). The CLHLS selected participants in a multistage, conducting surveys in both rural and urban regions of 23 out of 31 provinces in China, and covering approximately 85 % of the total population of those provinces. CLHLS is one of the most representative national age-related research databases in China and has been widely used (Xi et al., 2024; Zeng et al., 2017). The research ethics committees of Peking University (no. IRB00001052-13074) and all participants completed the informed consent before the survey.

This study uses two longitudinal datasets from CLHLS (i.e., the 2008–2018 wave and the 2011-2018 wave). We include three waves of data from CLHLS, covering a six-year follow-up period. The baseline survey of the current research was conducted in 2008 or 2011, and two subsequent follow-up surveys. For this study, the eligible participants only include that 1) 65 years or older at baseline; 2) have no cognitive impairment (MMSE <24) at baseline (Lou et al., 2023); 3) completion of the MMSE at follow-up; 4) have valid data for sociodemographic information, lifestyle, health conditions, and resilience measures for three waves (i.e., baseline, wave 1 and wave 2). The final sample consisted of 2,788 individuals in the main analysis and 1,070 in the sensitive analysis. A flowchart of the participants included in this study is illustrated in Figure 1.

**Figure 1.**
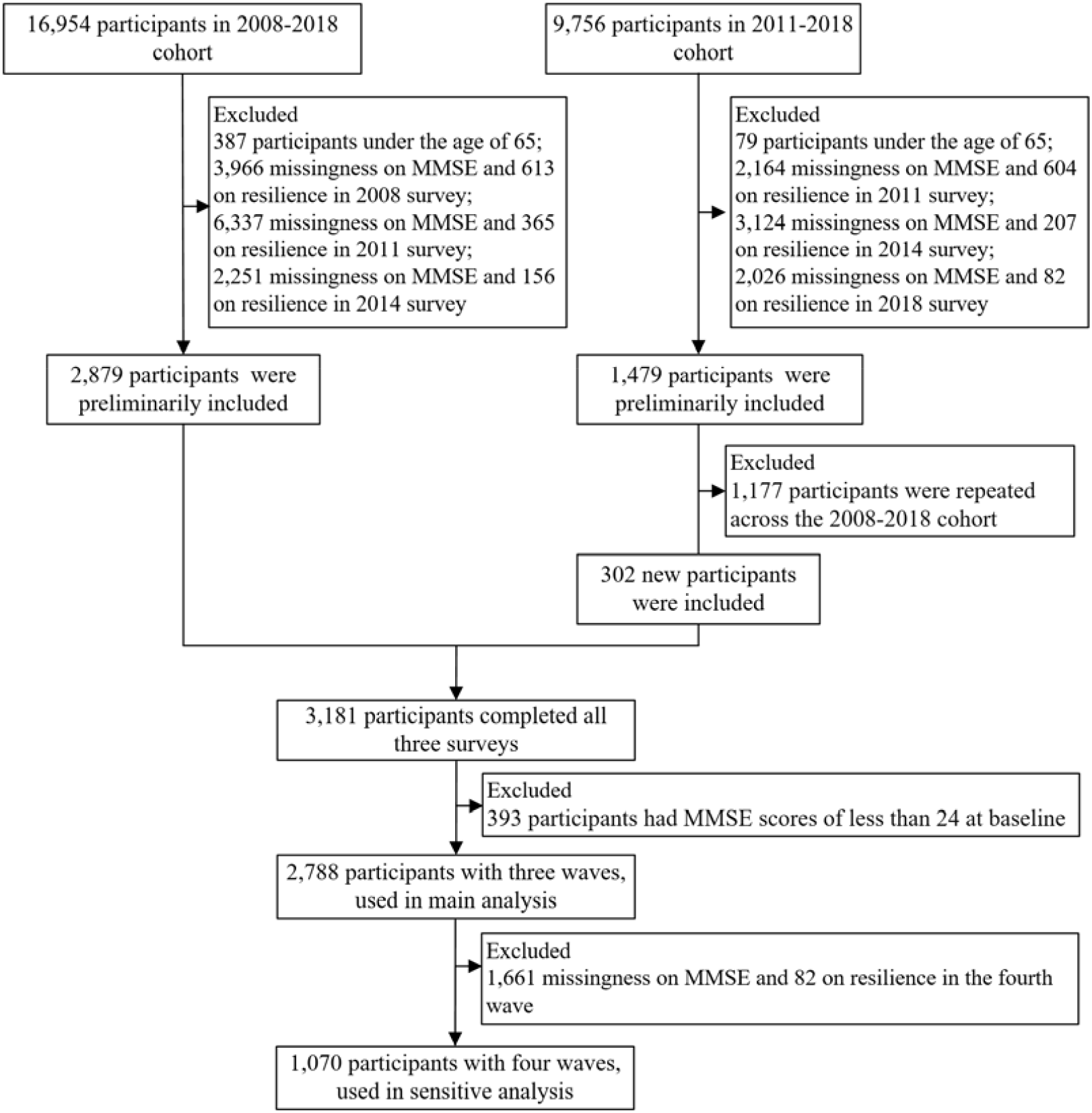
Flow diagram of participants included in the study.

## Measures

### Resilience Trajectories

Following a validated and published method, psychological resilience was measured using five items in the CLHLS dataset (Gu & Feng, 2018; Y. Yang & Wen, 2015):1)”Do you always look on the bright side of things?”, 2)”Can you make your own decisions concerning your personal affairs?”, 3)”Do you find that the older you get, the more useless you are, and have trouble doing anything?”, 4)”Do you often feel fearful or anxious?”, and 5)”Do you often feel lonely and isolated?”. The answers ranged from 1 (always) to 5 (never), and the total score ranged from 5 to 25, with reverse scoring applied to items 1 and 2. Higher scores indicated a higher level of psychological resilience, and the resilience scale has an acceptable value of internal consistency reliability (Cronbach’s alpha coefficient = 0.644) in this study. We identified three classes of resilience trajectories: the decreasing resilience group, the persistent middle resilience group, and the persistent high resilience group. A visual depiction of the trajectories and the detailed model fit metrics were shown in Figure 2 and Table S1.

**Figure 2.**
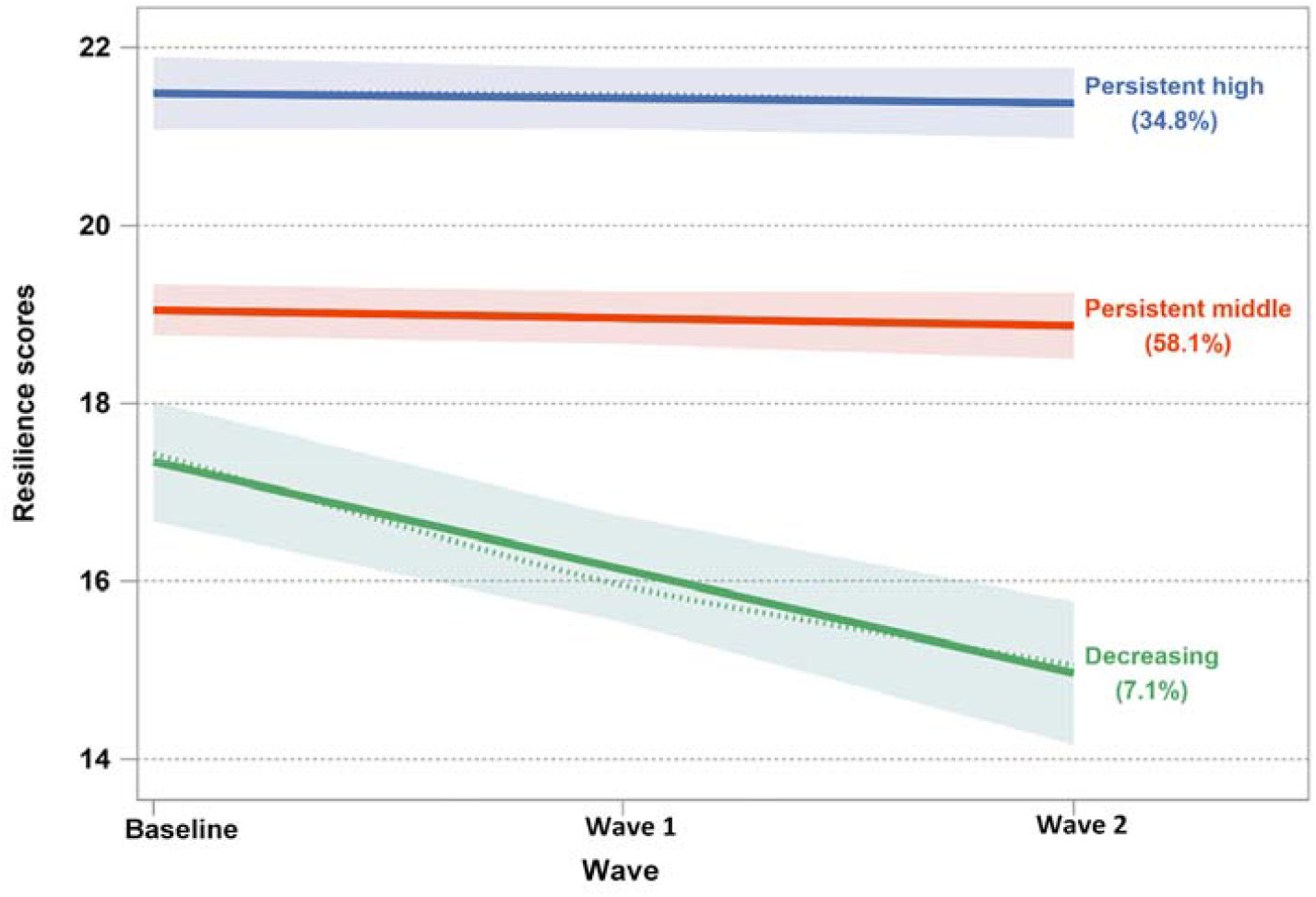
Psychological resilience trajectories in CLHLS cohort. The different colors of points represent the raw data points of participants in different resilience trajectories. The Y-axis represents psychological resilience scores, and the X-axis represents baseline and follow-up waves.

### Cognitive Impairment

Cognitive impairment was measured using the Chinese version of the Mini-Mental State Examination (MMSE) in the CLHLS dataset, which has been validated for reliability in previous studies among older adults (Xi et al., 2024; Yao et al., 2022). The test consists of 24 self-reported questions examining five cognitive dimensions and each question was scored as zero (incorrect or not answered) or one (correct), with a total score ranging from 0 to 30 and higher scores represent a better cognitive function. Previous studies reported the adequate reliability and validity of a cutoff point of 24 (Lou et al., 2023; Lv et al., 2019). Therefore, individuals with MMSE scores of <24 were identified as having cognitive impairment in this study. In the sensitivity analysis, we used a different cut-off of 18 MMSE scores for the definition of cognitive impairment (< 18 points) (Gao et al., 2017; Xi et al., 2024).

### Covariates

Sociodemographic characteristics, lifestyle and health conditions were chosen based on previous research findings at baseline. Sociodemographic variables included age, sex, region (east, central, west, or northeast), residence (rural or urban), educational level (years of education), marital status (married or otherwise), and occupation (agriculture, staff, or others). Lifestyle and health conditions comprised smoking and drinking status (yes or no), physical activity (yes or no), and the presence of hypertension (yes or no) and diabetes (yes or no).

### Statistical Analysis

Data were described as mean (SD) or median (IQR) for continuous variables, and frequency with percentage was used to describe categorical variables. Group-based trajectory modeling (GBTM) was used to identify possible trajectories of psychological resilience across the 3 waves (six years of follow-up). GBTM assumes that there are different trajectories among populations and uses maximum likelihood estimation to identify clusters of individuals with similar developmental trajectories (Huang et al., 2024; Nagin, 2005). To determine the best-fit number of resilience trajectories, we fitted models with 1 to 5 groups of trajectories and tested the significance of intercept, linear, and quadratic terms of time. Model fit was assessed using the Bayesian Information Criterion (BIC), where a lower BIC value indicates a better model fit. The average posterior probability (AvePP) was used to evaluate how well the model corresponds with the data, considering an AvePP of 0.7 or higher for each trajectory group as indicative of a good fit. Additionally, a higher relative entropy (Ek) value, approaching 1, was preferred for optimal model performance.

After identifying the trajectory clusters through GBTM, baseline characteristics were summarized based on the social isolation trajectory group and compared between participants using the chi-square test, ANOVA, or Kruskal-Wallis test, as appropriate. Cox proportional hazard models were used to estimate the hazard ratios (HRs) and 95% confidence intervals (CIs) for the associations between the psychological resilience trajectories and incident cognitive impairment using the persistent high resilience group as the reference. Model 1 was the unadjusted model, which included only resilience trajectories. Model 2 additionally adjusted for age, gender, region, residence, education level, marital status, occupation, smoking status, drinking status, and physical activity status. Model 3 was further adjusted for hypertension and diabetes based on Model 2. Furthermore, we used subgroup analyses to assess the potential difference in the associations between the psychological resilience trajectories and cognitive impairment by the lifestyle (smoking, drinking, and physical activity status) and health conditions (hypertension, diabetes).

We conducted a series of sensitivity analyses to assess the robustness of our findings. First, we repeated the Cox regression models using an MMSE score of 18 as the cut-off to define cognitive impairment (n=3,053). Second, we extended the follow-up period to include individuals who completed all four waves from 2008 to 2018 (covering a 10-year follow-up, n=1,070). We then repeated the GBTM and Cox regression to ensure the robustness of our results. All analyses were conducted using SAS 9.4 (SAS Institute, Cary, NC, USA), and two-sided *P*<0.05 was considered statistically significant.

## Results

### Sample characteristics

The baseline characteristics of participants based on psychological resilience trajectories are presented in Table 1. A total of 2,788 older adults were included in this study, the mean age of the study population at baseline was 79.4 (SD: 7.9) years, and the proportion of male participants was 53.2% (1483/2788).

**Table 1.** Sociodemographic characteristics of the participants at baseline.

| Characteristics | Total (n=2,788) | Resilience trajectory group |  |  | P-value <sup>a</sup> |
| --- | --- | --- | --- | --- | --- |
|  |  | Decreasing<br>(n=131) | Persistent middle<br>(n=1,808) | Persistent high<br>(n=849) |  |
| Age (years), mean (SD) | 79.4 (7.9) | 80.9 (7.7) | 79.7 (8.1) | 78.6 (7.4) | <0.001 |
| Sex, male, n (%) | 1,483 (53.2) | 32 (24.4) | 896 (49.6) | 555 (65.4) | <0.001 |
| Region, n (%) |  |  |  |  |  |
| East | 1,109 (39.8) | 41 (31.3) | 667 (36.9) | 401 (47.2) | <0.001 |
| Central | 723 (25.9) | 51 (38.9) | 525 (29.0) | 147 (17.3) |  |
| West | 764 (27.4) | 30 (22.9) | 513 (28.4) | 221 (26) |  |
| Northeast | 192 (6.9) | 9 (6.9) | 103 (5.7) | 80 (9.4) |  |
| Residence, n (%) |  |  |  |  |  |
| Urban | 1,484 (53.2) | 53 (40.5) | 894 (49.4) | 537 (63.3) | <0.001 |
| Rural | 1,304 (46.8) | 78 (59.5) | 914 (50.6) | 312 (36.7) |  |
| Education (years), median (IQR) | 2.0 (6.0) | 0 (2.0) | 1.0 (5.0) | 4.0 (7.0) | <0.001 |
| Married, n (%) | 1,468 (52.7) | 33 (25.2) | 889 (49.2) | 546 (64.3) | <0.001 |
| Occupation |  |  |  |  |  |
| Agriculture | 1,635 (58.6) | 97 (74.0) | 1,149 (63.6) | 389 (45.8) | <0.001 |
| Staff | 677 (24.3) | 10 (7.6) | 333 (18.4) | 334 (39.3) |  |
| Others | 476 (17.1) | 24 (18.3) | 326 (18.0) | 126 (14.8) |  |
| Smoking status, n (%) | 1,083 (38.9) | 28 (21.4) | 649 (35.9) | 406 (47.8) | <0.001 |
| Drinking status, n (%) | 936 (33.6) | 24 (18.3) | 577 (31.9) | 335 (39.5) | <0.001 |
| Physical activity, n (%) | 1,463 (52.5) | 48 (36.6) | 840 (46.5) | 575 (67.7) | <0.001 |
| Hypertension, n (%) | 1,196 (42.9) | 62 (47.3) | 780 (43.1) | 354 (41.7) | 0.451 |
| Diabetes, n (%) | 451 (16.2) | 24 (18.3) | 264 (14.6) | 163 (19.2) | 0.009 |

Note: <sup>a</sup> P-value was based on the chi-square test, ANOVA, or the Kruskal-Wallis test where appropriate.

### Trajectories of psychological resilience

In trajectory analyses, we identified three distinct psychological resilience trajectories over a 6-year follow-up period (Figure 2). The results of GBTM showed that both the two-trajectory model and the three-trajectory model have an AvePP index greater than 0.7. Taking into account the BIC, AvePP index, and Ek values, we selected the three-trajectory model as the best-fitting model (Table S1).

Figure 2 illustrated the characteristics of the three trajectories we identified. The first is characterized by decreasing resilience scores (decreasing resilience group, 131 [7.1%]), with initial resilience scores being lowest at baseline (mean, 16.9; SD, 2.4) and then decreasing to nearly 3 points at wave 1 and wave 2. The second trajectory is characterized by persistent middle resilience scores (persistent middle resilience group, 1,808 [58.1%]), and psychological resilience scores were close to 18.8 points at each time point. The third trajectory is characterized by persistently high psychological resilience scores throughout the follow-up period (persistent high resilience group, 849 [34.8%]), with scores consistently around 22 points at each time point.

The characteristics of the participants in the resilience trajectory group were described in Table 1. Compared with those in the persistent middle resilience group or high resilience group, those in the decreasing resilience group were more likely to be older, live in rural areas, less educated, unmarried, work in agriculture, less physical activity, non-drinkers and non-smokers, and without diabetes (*P*-value <0.05).

### Resilience Trajectories and Cognitive Impairment

Of the 2,778 participants in this study, 571 individuals (20.6%) developed cognitive impairments after 6 years of follow-up. As shown in Figure 3, compared to the persistent high resilience group, people with persistent middle resilience (HR = 1.94, 95%CI = 1.56-2.41) and decreasing resilience (HR = 4.39, 95%CI = 3.20-6.02) were significantly associated with a higher risk of developing cognitive impairment in crude model (Model 1). These associations remained significant after adjusting for the sociodemographic and lifestyle at baseline (Model 2). After adding health conditions to the model, persistent middle resilience (HR = 1.43, 95%CI = 1.14-1.79) and decreasing resilience (HR = 2.46, 95%CI = 1.76-3.43) remained consistently associated with higher odds of having cognitive impairment (Table S2).

**Figure 3.**
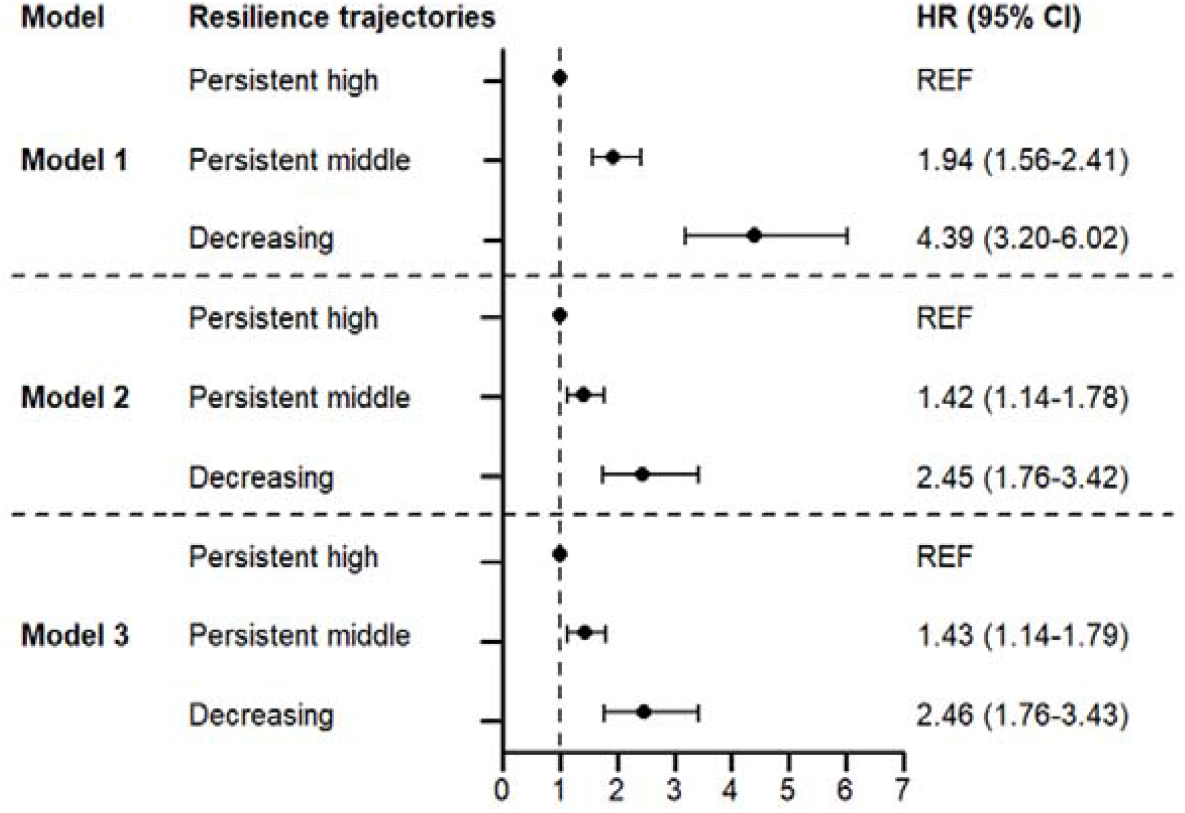
Associations between psychological resilience trajectories and cognitive impairment. The persistent high resilience group was as a reference group. Model 1 was a crude model; model 2 additionally adjusted for age, gender, region, residence, education level, marital status, occupation, smoking status, drinking status, and physical activity status; model 3 further adjusted for health conditions based on model 2. Abbreviations: HR, hazard ratios; 95% CI, 95% Confidence Intervals; REF, reference.

### Subgroup analyses by lifestyle and health conditions

We conducted a series of subgroup analyses to explore potential differences among participants stratified by lifestyle and health conditions (Table 2). Compared to participants with healthy lifestyle conditions (non-smokers, HR = 0.35, 95%CI = 0.23-0.52; non-drinkers, HR =0.37, 95%CI = 0.25-0.55), the persistent high trajectory of resilience had a lower risk of cognitive impairment for those who with unhealthy lifestyle conditions (smokers, HR = 0.52, 95%CI = 0.25-1.06; drinkers, HR = 0.47, 95%CI = 0.23-0.98). Among exercisers, participants with persistent high resilience (HR = 0.39, 95%CI = 0.24-0.63) had greater protection against cognitive impairment compared to non-exercisers (HR = 0.42, 95%CI = 0.26-0.70). When each chronic disease was analyzed individually, compared with older adults who had hypertension (HR = 0.34, 95%CI = 0.20-0.56) or diabetes (HR = 0.23, 95%CI = 0.10-0.54), the persistent high resilience trajectories posed a lower risk of cognitive impairment for those who did not have chronic diseases (without hypertension, HR = 0.41, 95%CI = 0.31-0.76; without diabetes, HR = 0.45, 95%CI = 0.31-0.66). A similar association was observed in the persistent middle resilience trajectory group, as detailed in Table 2.

**Table 2.** The association between resilience trajectories and cognitive impairment, stratified by lifestyle and health conditions.

| Resilience trajectories | HR (95% CI) | HR (95% CI) |
| --- | --- | --- |
| **Smoking status** | **Non-smoker** | **Smoker** |
| Decreasing | 1.00 (ref.) | 1.00 (ref.) |
| Persistent high | **0.35 (0.23-0.52)** | 0.52 (0.25-1.06) |
| Persistent middle | **0.59 (0.43-0.80)** | 0.57 (0.30-1.11) |
| **Drinking status** | **Non-drinkers** | **Drinker** |
| Decreasing | 1.00 (ref.) | 1.00 (ref.) |
| Persistent high | **0.37 (0.25-0.55)** | **0.47 (0.23-0.98)** |
| Persistent middle | **0.59 (0.43-0.80)** | 0.57 (0.30-1.09) |
| **Physical activity status** | **Exerciser** | **Non-exerciser** |
| Decreasing | 1.00 (ref.) | 1.00 (ref.) |
| Persistent high | **0.39 (0.24-0.63)** | **0.42 (0.26-0.70)** |
| Persistent middle | **0.53 (0.38-0.76)** | **0.60 (0.38-0.94)** |
| **Hypertension** | **Without hypertension** | **With hypertension** |
| Decreasing | 1.00 (ref.) | 1.00 (ref.) |
| Persistent high | **0.41 (0.31-0.76)** | **0.34 (0.20-0.56)** |
| Persistent middle | **0.67 (0.46-0.98)** | **0.50 (0.33-0.74)** |
| **Diabetes** | **Without diabetes** | **With diabetes** |
| Decreasing | 1.00 (ref.) | 1.00 (ref.) |
| Persistent high | **0.45 (0.31-0.66)** | **0.23 (0.10-0.54)** |
| Persistent middle | **0.63 (0.46-0.85)** | **0.41 (0.21-0.79)** |

Note: All models adjusted for age, gender, region, residence, education level, marital status, and occupation. Health-related factors (i.e., smoking status, drinking status, physical activity status, hypertension, and diabetes) were sequentially included as stratified variables, while the other four factors were controlled as covariates in the models.

### Sensitive analysis

The robustness of the results was confirmed by sensitivity analyses. When older adults with an MMSE score of less than 18 were considered cognitive impaired, 7.0% (215/3,053) of them developed cognitive impairment during the 6 years of follow-up. The persistent middle resilience (HR = 3.14, 95% CI = 1.87-5.29) and decreasing resilience (HR = 5.00, 95% CI = 2.64-9.48) were associated with a higher risk of cognitive impairment (Table S3). We further included individuals who completed four waves of the surveys (10 years of follow-up, n=1,070) for sensitive analysis. We obtained similar results that identified a three-trajectory model with similar characteristics of each cluster, namely persistent high resilience trajectory, persistent middle resilience trajectory, and decreasing resilience trajectory (Figure S1). Of these 1,070 participants, 146 (13.6%) developed cognitive impairment. Similarly, in the fully adjusted models, persistent middle resilience (HR = 2.88, 95% CI = 1.14-1.79) and decreasing resilience (HR = 4.46, 95% CI = 1.36-14.68) were associated with a higher risk of cognitive impairment (Table S4).

## Discussion

In this cohort study, three distinct psychological resilience trajectories were identified for Chinese older adults over a 6-year follow-up: persistent high resilience trajectory group, persistent middle resilience trajectory group, and decreasing resilience trajectory group. Participants in the persistent middle and decreasing resilience trajectory group have a higher risk of developing cognitive impairment compared to those in the persistent high group, with decreasing resilience trajectory being particularly harmful. This association remains consistently significant after adjusting for sociodemographic factors, lifestyle and health conditions. This association seemed to be stronger among nonsmokers, nondrinkers, exercisers, and older adults with hypertension and diabetes. Our findings highlighted the importance of protecting against declines in psychological resilience from a long-term perspective to prevent cognitive decline.

Psychological resilience in older adults can be defined as the capacity to maintain health, well-being, and life satisfaction in the face of challenges (Kiosses & Sachs-Ericsson, 2020; Manning, 2015). High resilience has been found to be associated with positive outcomes, including successful aging, lower depression, better self-perceptions and lifestyle behaviors, and longevity (Angevaare et al., 2020; Smith & Hollinger-Smith, 2015). In this study, we found that psychological resilience is generally a stable trait among older adults in China, supporting the findings of previous research (Taylor & Carr, 2021). While most participants maintained stable high or middle levels of psychological resilience, approximately 7% exhibited a decline over time, highlighting the importance of monitoring psychological resilience changes among older adults. Additionally, the trajectory characteristics show that older adults in the decreasing resilience group not only experienced a decline in resilience over the follow-up period, but also had lower baseline resilience scores compared to those in the persistent high and persistent middle groups. This may suggest that people with lower psychological resilience might be more prone to experiencing a downward trend. Consistently to previous studies (Jiang et al., 2024; MacLeod et al., 2016;), this group is also more likely to be older, female, rural, less educated, and unmarried. Further studies are therefore needed to better understand the nature of different trajectories and to explore factors that influence resilience trajectories, with the aim of developing effective interventions to protect against declining psychological resilience.

Similar to previous studies showing that higher resilience is associated with better cognitive function and slower cognitive decline (Jiang et al., 2024; Kiosses & Sachs-Ericsson, 2020; Lou et al., 2023; J. S. Yang et al., 2021), findings of this study underscore the association between resilience trajectories and cognitive impairment among older adults. Compared with those in the persistent high resilience group, those in the persistent middle or declining group were at higher risk of cognitive impairment, with the risk being greater for those in the declining resilience group. Previous research has shown that psychological resilience is associated with various positive health outcomes, such as improved quality of life, better mental health, and increased physical activity (Moore et al., 2015; Pardeller et al., 2020), all of which may further be linked to cognitive function. Existing research has observed that individuals with lower resilience capacity experienced greater amyloid-related cognitive decline over time, but the mechanisms underlying resilience against amyloid pathology remain unclear (Wolf et al., 2019). Although several etiological hypotheses, including brain or cognitive reserve, vascular and stress hypotheses, have been proposed to explain the relationship between psychological resilience and cognitive impairment, the exact mechanisms are still unknown (Giovacchini et al., 2019; Lou et al., 2023). The persistent high resilience group likely benefits from these protective psychological mechanisms and maintains cognitive health, while further studies are needed to investigate these underlying mechanisms, particularly from the perspective of long-term changes in psychological resilience.

Differences in the associations between resilience trajectories and cognitive impairment were found between varied lifestyle and health conditions groups in this research. Previous research found that the association between resilience and cognitive function was moderated by body composition, such as inflammatory status (Jung et al., 2021). We have added evidence regarding lifestyle and chronic diseases, which are crucial modifiable factors in the daily lives of older adults. Compared with smokers and drinkers, participants with a healthy lifestyle (i.e. non-smokers, non-drinkers) who had a persistent high resilience trajectory were less likely to develop cognitive impairment, possibly due to the greater detrimental impact of smoking and drinking on cognitive health (Livingston et al., 2020). Aligned with previous studies that found a positive relationship between physical activity and resilience in older adults (Toth et al., 2024; Wermelinger et al., 2018), our study supports that higher resilience provides better protection for cognitive function compared to non-exercisers. This may be because physical activity enhances neuroplasticity and cognitive reserve, thereby reducing the susceptibility to cognitive decline (Kivipelto et al., 2018; Sofi et al., 2011). Furthermore, we found that those with persistent high resilience trajectory is associated with a lower risk of developing cognitive impairment, particularly in older adults with chronic diseases, indicating that resilience has a more pronounced protective effect in those with health challenges. This finding aligns with previous research, which has shown that greater psychological resilience is associated with better physical functioning, lower self-reported disability, better mental quality of life, and lower likelihood of frailty among older individuals with a history of type 2 diabetes mellitus (Olson et al., 2023; Pesantes et al., 2015). Enhancing psychological resilience in individuals with health conditions is crucial.

Interventions to promote or maintain high psychological resilience and prevent its decline are crucial for better cognitive health and ensuring successful aging. Current interventions that highlight optimism and positive emotions may be particularly effective in building resilience, such as cognitive behavioral therapy and mindfulness activities aimed at enhancing happiness (Gooding et al., 2012; MacLeod et al., 2016; Southwick et al., 2014). Additionally, an innovative approach focuses on enhancing gratitude, challenging aging stereotypes, and encouraging older adults to engage in value-based activities (Treichler et al., 2020). Incorporating physical activity, increasing social support, and improving social relationships are also suggested as effective alternative methods (Izquierdo & Fiatarone Singh, 2023; Peeters et al., 2023). The findings of our study can serve as supplementary evidence, emphasizing that, alongside specific interventions, healthy lifestyles should be valued, and targeted strategies for health conditions should not be overlooked.

The present study had several limitations. First, although our resilience was based on previous research, it was not a standardized scale or instrument, and it relies on self-report and inevitably has potential information bias. Second, we used the more convenient MMSE rather than neuropsychological tests to assess cognitive functioning, and the MMSE has limited sensitivity to changes in cognitive feats in some populations. Third, despite these findings providing valuable insights, there are possibilities of reverse causality due to the observational nature of the study. Finally, some correlates of cognitive impairment, such as social support and loneliness, were not included in our analysis due to the limitations of the original survey study. Future research should aim to include as many confounding variables as possible to thoroughly investigate the independent effects of resilience trajectories on cognitive impairment as well as the underlying causes and mechanisms.

## Conclusion

In summary, the present study identified three trajectories of psychological resilience among older adults in China: the majority clustered into the persistent high resilience and persistent middle resilience groups, while a smaller proportion were in the decreasing resilience group. Compared with those in the persistent high resilience group, both the persistent middle and decreasing resilience groups were significantly associated with an increased risk of developing cognitive impairment, with slight variations observed in subgroups stratified by lifestyle and health conditions. To our knowledge, this is the first study to examine the association between resilience trajectories and cognitive impairment. Our findings suggest that maintaining and improving psychological resilience over time is important for cognitive health, particularly for those with unhealthy lifestyles and health conditions.

## Data Availability

This study was not preregistered. The data supporting the findings of this study are from the publicly available CLHLS data set, which is available from their official website (https://opendata.pku.edu.cn/dataverse/CHADS). The details of analytic methods can be made available upon reasonable request.

## Supplementary Material

Supplementary data are available at online.

## References

Angevaare, M. J., Roberts, J., van Hout, H. P. J., Joling, K. J., Smalbrugge, M., Schoonmade, L. J., … Hertogh, C. M. P. M. (2020). Resilience in older persons: A systematic review of the conceptual literature. Ageing Research Reviews, 63, 101144. 10.1016/j.arr.2020.101144

Anstey, K. J., von Sanden, C., Salim, A., & O’Kearney, R. (2007). Smoking as a risk factor for dementia and cognitive decline: a meta-analysis of prospective studies. American Journal of Epidemiology, 166(4), 367–378. 10.1093/aje/kwm116

Feigin, V. L., Nichols, E., Alam, T., Bannick, M. S., Beghi, E., Blake, N., Vos, T. (2019). Global, regional, and national burden of neurological disorders, 1990–2016: a systematic analysis for the Global Burden of Disease Study 2016. The Lancet Neurology, 18(5), 459–480. 10.1016/S1474-4422(18)30499-X

Franks, K. H., Bransby, L., Cribb, L., Buckley, R., Yassi, N., Chong, T. T. J., Pase, M. P. (2023). Associations of Perceived Stress and Psychological Resilience With Cognition and a Modifiable Dementia Risk Score in Middle-Aged Adults. The Journals of Gerontology. Series B, Psychological Sciences and Social Sciences, 78(12), 1992–2000. 10.1093/geronb/gbad131

Franks, K. H., Rowsthorn, E., Bransby, L., Lim, Y. Y., Chong, T. T. J., & Pase, M. P. (2023). Association of Self-Reported Psychological Stress with Cognitive Decline: A Systematic Review. Neuropsychology Review, 33(4), 856–870.10.1007/s11065-022-09567-y

Gallardo-Gómez, D., Del Pozo-Cruz, J., Noetel, M., Álvarez-Barbosa, F., Alfonso-Rosa, R. M., & Del Pozo Cruz, B. (2022). Optimal dose and type of exercise to improve cognitive function in older adults: A systematic review and bayesian model-based network meta-analysis of RCTs. Ageing Research Reviews, 76, 101591. 10.1016/j.arr.2022.101591

Gao, M., Kuang, W., Qiu, P., Wang, H., Lv, X., & Yang, M. (2017). The time trends of cognitive impairment incidence among older Chinese people in the community: based on the CLHLS cohorts from 1998 to 2014. Age and Ageing, 46(5), 787–793. 10.1093/ageing/afx038

Giovacchini, G., Giovannini, E., Borsò, E., Lazzeri, P., Riondato, M., Leoncini, R., Ciarmiello, A. (2019). The brain cognitive reserve hypothesis: A review with emphasis on the contribution of nuclear medicine neuroimaging techniques. Journal of Cellular Physiology, 234(9), 14865–14872. 10.1002/jcp.28308

Gooding, P. A., Hurst, A., Johnson, J., & Tarrier, N. (2012). Psychological resilience in young and older adults. International Journal of Geriatric Psychiatry, 27(3), 262–270. 10.1002/gps.2712

Gu, D., & Feng, Q. (2018). Psychological Resilience of Chinese Centenarians and Its Associations With Survival and Health: A Fuzzy Set Analysis. The Journals of Gerontology. Series B, Psychological Sciences and Social Sciences, 73(5), 880–889.10.1093/geronb/gbw071

Harvanek, Z. M., Fogelman, N., Xu, K., & Sinha, R. (2021). Psychological and biological resilience modulates the effects of stress on epigenetic aging. Translational Psychiatry, 11(1), 601. 10.1038/s41398-021-01735-7

Huang, Y., Zhu, X., Liu, X., & Li, J. (2024). Loneliness Trajectories Predict Risks of Cardiovascular Diseases in Chinese Middle-Aged and Older Adults. The Journals of Gerontology. Series B, Psychological Sciences and Social Sciences, 79(5). 10.1093/geronb/gbae018

Izquierdo, M., & Fiatarone Singh, M. (2023). Promoting resilience in the face of ageing and disease: The central role of exercise and physical activity. Ageing Research Reviews, 88, 101940. 10.1016/j.arr.2023.101940

Jia, J., Zhao, T., Liu, Z., Liang, Y., Li, F., Li, Y., … Cummings, J. (2023). Association between healthy lifestyle and memory decline in older adults: 10 year, population based, prospective cohort study. BMJ (Clinical Research ed.), 380, e072691. 10.1136/bmj-2022-072691

Jiang, G.-Q., He, Y.-K., Li, T.-F., Qin, Q.-R., Wang, D.-N., Huang, F., … Li, J. (2024). Association of psychological resilience and cognitive function in older adults: Based on the Ma’ anshan Healthy Aging Cohort Study. Archives of Gerontology and Geriatrics, 116, 105166. 10.1016/j.archger.2023.105166

Jung, S. J., Lee, G. B., Nishimi, K., Chibnik, L., Koenen, K. C., & Kim, H. C. (2021). Association between psychological resilience and cognitive function in older adults: effect modification by inflammatory status. GeroScience, 43(6), 2749–2760. 10.1007/s11357-021-00406-1

Kiosses, D. N., & Sachs-Ericsson, N. (2020). Increasing resilience in older adults. International Psychogeriatrics, 32(2), 157–159. 10.1017/S1041610220000046

Kivipelto, M., Mangialasche, F., & Ngandu, T. (2018). Lifestyle interventions to prevent cognitive impairment, dementia and Alzheimer disease. Nature Reviews. Neurology, 14(11), 653–666. 10.1038/s41582-018-0070-3

Linnemann, P., Wellmann, J., Berger, K., & Teismann, H. (2020). Effects of age on trait resilience in a population-based cohort and two patient cohorts. Journal of Psychosomatic Research, 136, 110170. 10.1016/j.jpsychores.2020.110170

Livingston, G., Huntley, J., Sommerlad, A., Ames, D., Ballard, C., Banerjee, S., Mukadam, N. (2020). Dementia prevention, intervention, and care: 2020 report of the Lancet Commission. Lancet, 396(10248), 413–446. 10.1016/s0140-6736(20)30367-6

Lou, Y., Irakoze, S., Huang, S., You, Q., Wang, S., Xu, M., Cao, S. (2023). Association of social participation and psychological resilience with adverse cognitive outcomes among older Chinese adults: A national longitudinal study. Journal of Affective Disorders, 327, 54–63. 10.1016/j.jad.2023.01.112

Lv, X., Li, W., Ma, Y., Chen, H., Zeng, Y., Yu, X., Wang, H. (2019). Cognitive decline and mortality among community-dwelling Chinese older people. BMC Medicine, 17(1), 63. 10.1186/s12916-019-1295-8

Machado, A., Herrera, A. J., de Pablos, R. M., Espinosa-Oliva, A. M., Sarmiento, M., Ayala, A., … Cano, J. (2014). Chronic stress as a risk factor for Alzheimer’s disease. Reviews In the Neurosciences, 25(6), 785–804. 10.1515/revneuro-2014-0035

MacLeod, S., Musich, S., Hawkins, K., Alsgaard, K., & Wicker, E. R. (2016). The impact of resilience among older adults. Geriatric Nursing (New York, N.Y.), 37(4), 266–272.10.1016/j.gerinurse.2016.02.014

Manning, L. K. (2015). Resilience and Aging: From Conceptual Understandings to Opportunities for Enhancement. The Gerontologist, 55(4), 703–704. 10.1093/geront/gnv086

McDaniel, J. T., Hascup, E. R., Hascup, K. N., Trivedi, M., Henson, H., Rados, R., … Frick, K. (2022). Psychological Resilience and Cognitive Function Among Older Military Veterans. Gerontology & Geriatric Medicine, 8, 23337214221081363. 10.1177/23337214221081363

Moore, R. C., Eyler, L. T., Mausbach, B. T., Zlatar, Z. Z., Thompson, W. K., Peavy, G., Jeste, D. V. (2015). Complex interplay between health and successful aging: role of perceived stress, resilience, and social support. The American Journal of Geriatric Psychiatry : Official Journal of the American Association For Geriatric Psychiatry, 23(6), 622–632. 10.1016/j.jagp.2014.08.004

Nagin, D. S. (2005). Group-Based Modeling of Development: Harvard University Press.

Olson, K. L., Howard, M., McCaffery, J. M., Dutton, G. R., Espeland, M. A., Simpson, F. R., Wing, R. R. (2023). Psychological resilience in older adults with type 2 diabetes from the Look AHEAD Trial. Journal of the American Geriatrics Society, 71(1), 206–213.10.1111/jgs.17986

Pardeller, S., Kemmler, G., Hoertnagl, C. M., & Hofer, A. (2020). Associations between resilience and quality of life in patients experiencing a depressive episode. Psychiatry Research, 292, 113353. 10.1016/j.psychres.2020.113353

Peeters, G., Kok, A., de Bruin, S. R., van Campen, C., Graff, M., Nieuwboer, M., Olde Rikkert, M. (2023). Supporting Resilience of Older Adults with Cognitive Decline Requires a Multi-Level System Approach. Gerontology, 69(7), 866–874. 10.1159/000529337

Pesantes, M. A., Lazo-Porras, M., Abu Dabrh, A. M., Ávila-Ramírez, J. R., Caycho, M., Villamonte, G. Y., … Miranda, J. J. (2015). Resilience in Vulnerable Populations With Type 2 Diabetes Mellitus and Hypertension: A Systematic Review and Meta-analysis. The Canadian Journal of Cardiology, 31(9), 1180–1188. 10.1016/j.cjca.2015.06.003

Smith, J. L., & Hollinger-Smith, L. (2015). Savoring, resilience, and psychological well-being in older adults. Aging & Mental Health, 19(3), 192–200. 10.1080/13607863.2014.986647

Sofi, F., Valecchi, D., Bacci, D., Abbate, R., Gensini, G. F., Casini, A., & Macchi, C. (2011). Physical activity and risk of cognitive decline: a meta-analysis of prospective studies. Journal of Internal Medicine, 269(1), 107–117. 10.1111/j.1365-2796.2010.02281.x

Southwick, S. M., Bonanno, G. A., Masten, A. S., Panter-Brick, C., & Yehuda, R. (2014). Resilience definitions, theory, and challenges: interdisciplinary perspectives. European Journal of Psychotraumatology, 5. 10.3402/ejpt.v5.25338

Taylor, M. G., & Carr, D. (2021). Psychological Resilience and Health Among Older Adults: A Comparison of Personal Resources. The Journals of Gerontology. Series B, Psychological Sciences and Social Sciences, 76(6), 1241–1250. 10.1093/geronb/gbaa116

Topiwala, A., Allan, C. L., Valkanova, V., Zsoldos, E., Filippini, N., Sexton, C., Ebmeier, K. P. (2017). Moderate alcohol consumption as risk factor for adverse brain outcomes and cognitive decline: longitudinal cohort study. BMJ (Clinical Research ed.), 357, j2353. 10.1136/bmj.j2353

Toth, E. E., Ihász, F., Ruíz-Barquín, R., & Szabo, A. (2024). Physical Activity and Psychological Resilience in Older Adults: A Systematic Review of the Literature. Journal of Aging and Physical Activity, 32(2), 276–286. 10.1123/japa.2022-0427

Treichler, E. B. H., Glorioso, D., Lee, E. E., Wu, T.-C., Tu, X. M., Daly, R., Jeste, D. V. (2020). A pragmatic trial of a group intervention in senior housing communities to increase resilience. International Psychogeriatrics, 32(2), 173–182. 10.1017/S1041610219002096

Ungar, M., & Theron, L. (2020). Resilience and mental health: how multisystemic processes contribute to positive outcomes. The Lancet. Psychiatry, 7(5), 441–448. 10.1016/S2215-0366(19)30434-1

Vassilaki, M., Aakre, J. A., Cha, R. H., Kremers, W. K., St Sauver, J. L., Mielke, M. M., Roberts, R. O. (2015). Multimorbidity and Risk of Mild Cognitive Impairment. Journal of the American Geriatrics Society, 63(9), 1783–1790. 10.1111/jgs.13612

Wermelinger Ávila, M. P., Corrêa, J. C., Lucchetti, A. L. G., & Lucchetti, G. (2018). The Role of Physical Activity in the Association Between Resilience and Mental Health in Older Adults. Journal of Aging and Physical Activity, 26(2), 248–253. 10.1123/japa.2016-0332

Wolf, D., Fischer, F. U., & Fellgiebel, A. (2019). Impact of Resilience on the Association Between Amyloid-β and Longitudinal Cognitive Decline in Cognitively Healthy Older Adults. Journal of Alzheimer’s Disease : JAD, 70(2), 361–370. 10.3233/JAD-190370

Xi, D., Liu, L., Zhang, M., Huang, C., Burkart, K. G., Ebi, K., … Ji, J. S. (2024). Risk factors associated with heatwave mortality in Chinese adults over 65 years. Nature Medicine, 30(5), 1489–1498. 10.1038/s41591-024-02880-4

Yang, J. S., Jeon, Y. J., Lee, G. B., Kim, H. C., & Jung, S. J. (2021). The association between psychological resilience and cognitive function in longitudinal data: Results from the community follow-up survey. Journal of Affective Disorders, 290, 109–116. 10.1016/j.jad.2021.04.062

Yang, Y., & Wen, M. (2015). Psychological Resilience and the Onset of Activity of Daily Living Disability Among Older Adults in China: A Nationwide Longitudinal Analysis. The Journals of Gerontology. Series B, Psychological Sciences and Social Sciences, 70(3), 470–480. 10.1093/geronb/gbu068

Yao, Y., Lv, X., Qiu, C., Li, J., Wu, X., Zhang, H., Zeng, Y. (2022). The effect of China’s Clean Air Act on cognitive function in older adults: a population-based, quasi-experimental study. The Lancet. Healthy Longevity, 3(2). 10.1016/S2666-7568(22)00004-6

Yi, Z. (2008). Introduction to the Chinese Longitudinal Healthy Longevity Survey (CLHLS). In Z. Yi, D. L. Poston, D.A. Vlosky & D. Gu (Eds.), Healthy Longevity in China: Demographic, Socioeconomic, and Psychological Dimensions (pp. 23–38). Dordrecht: Springer Netherlands.

Zeng, Y. (2012). Towards Deeper Research and Better Policy for Healthy Aging --Using the Unique Data of Chinese Longitudinal Healthy Longevity Survey. China Economic Journal, 5(2-3), 131–149. 10.1080/17538963.2013.764677

Zeng, Y., Feng, Q., Hesketh, T., Christensen, K., & Vaupel, J. W. (2017). Survival, disabilities in activities of daily living, and physical and cognitive functioning among the oldest-old in China: a cohort study. Lancet (London, England), 389(10079), 1619–1629. 10.1016/S0140-6736(17)30548-2

